# Damage to serotonergic and opioid networks relates to post-stroke epilepsy after thrombectomy

**DOI:** 10.64898/2026.04.27.26351878

**Authors:** Benedikt Frey, Joachim Gruber, Philipp Koch, Hannes Deutschmann, Christian Enzinger, Jan Feldheim, Laura Hartmann, Raimund Helbok, Tom Hornberger, Markus Kneihsl, Daniela Pinter, Fanny Quandt, Stefan Ropele, Michael Sonnberger, Götz Thomalla, Thomas Gattringer, Robert Schulz

## Abstract

Post-stroke epilepsy (PSE) is a clinically relevant complication after ischemic stroke. While lesion location and established clinical risk factors contribute to PSE risk, the role of lesion-induced disruption of neurotransmitter-specific brain networks remains unclear. We retrospectively analyzed 251 patients with acute large-vessel occlusion ischemic stroke treated with mechanical thrombectomy. Binary lesion masks were embedded into normative neurotransmitter-informed structural connectomes derived from PET-based receptor and transporter density maps, yielding damage scores for 19 neurotransmitter systems and a global measure of structural disconnection. Partial least squares (PLS) regression was used for feature selection, followed by multivariable logistic regression adjusted for age, sex, and SeLECT. As a secondary internal resampling analysis, elastic-net logistic regression with repeated stratified cross-validation was used to assess whether the identified pattern remained informative under regularization. Twenty-six patients (10.4%) developed PSE. PLS identified a neurochemical signature dominated by serotonergic and µ-opioid–informed systems. In adjusted models, damage to 5-HT1a, 5-HT2a, and µ-opioid networks showed the strongest and most robust associations with PSE, independent of clinical predictors and global structural disconnection. Cholinergic network measures showed weaker and less consistent effects. In repeated internal cross-validation, the same networks were selected more consistently and were associated with higher discrimination than the clinical base model. Lesion-induced disruption of specific neurotransmitter-informed structural networks, most robustly serotonergic and µ-opioid systems, is associated with PSE and showed incremental signal in internal cross-validation beyond established clinical risk factors. These findings provide a mechanistically interpretable extension to existing PSE risk models.

## Introduction

Post-stroke epilepsy (PSE) is one of the most frequent and clinically relevant complications after ischemic stroke^1,2^. This condition affects up to 10 % of stroke survivors and contributes substantially to long-term morbidity and reduced quality of life. Younger patients with large cortical infarctions, watershed strokes, early seizures, and severe initial deficits are at a particular risk for PSE^3-7^. Recent research has provided novel insights into possible mechanisms of PSE at the systems level, with inconclusive results regarding specific locations that would lead to an increased risk^8,9^, but converging evidence that specific functional brain networks comprising cerebellar regions and the basal ganglia could be involved in PSE development^10^.

Despite these advances, current models are still far from being complete. A critical limitation, particularly regarding the neuropathophysiological aspects of neurotransmitter involvement in PSE^11^, is that the neurochemical component has been neglected in neuroimaging studies of PSE to date. PSE can be viewed as a disturbance of the excitatory-inhibitory balance. Gamma-aminobutyric acid (GABA), the primary inhibitory neurotransmitter, provides tonic inhibition through extrasynaptic GABA^A^ receptors. After a stroke, loss or dysfunction of these receptors reduces inhibitory tone, facilitating network hyperexcitability and seizure generation^11-13^. Conversely, glutamate, the major excitatory neurotransmitter, accumulates after ischemia, activating NMDA receptors and driving excitotoxic cascades, neuronal death, and synaptic reorganization^14-16^. However, the extent to which NMDA activation promotes PSE remains a topic of debate^17^. Serotonin has also been shown to modulate excitability and synaptic synchronization^18^. Animal studies suggest that increased extracellular serotonin, e.g., through SSRIs or precursors such as 5-hydroxytryptophan (5-HT), can raise seizure thresholds, whereas serotonin depletion can lower them^19,20^. However, serotonin effects on epileptic excitability remain complex with dosage, receptor, and regional specificity^18^. Acetylcholine has also been linked to cortical excitability, for instance, via muscarine M1 receptors, which can facilitate network synchronization and promote epileptic activity^21-23^. Finally, histamine^24^, cannabinoid^25^, and opioid systems^26^ have been equally linked to neural excitability or epileptogenesis. Whether incorporating neurochemical network vulnerability improves risk stratification beyond established clinical models remains unknown.

We hypothesize that integrating a neurochemical layer of analysis into neuroimaging can provide a more comprehensive understanding of the pathophysiology of PSE. This has recently been demonstrated for stroke deficits and global outcome by two independent studies, which extended their brain network analyses by normative neurochemical data on neurotransmitter (NT) receptors and transporters^27^ into their network analyses^28,29^. For instance, the disruption of dopaminergic brain networks, particularly characterized by the expression of the dopamine transporters, has been recently linked to a worse overall outcome after stroke^29^.

Given the central role of neurotransmitters in PSE, we hypothesized that PSE would not only associate with clinical variables, lesion characteristics, and structural disconnection, but that NT-informed network disruption, potentially within GABAergic, glutamatergic, serotonergic, and cholinergic systems, may provide additional explanatory value for established models of PSE occurrence in patients with large-vessel occlusion treated with mechanical thrombectomy.

## Methods and Materials

### Participants

Patients with acute large-vessel occlusion (LVO) stroke who underwent mechanical thrombectomy (MT) at the Department of Neurology 1, Neuromed Campus, Kepler University Hospital Linz, and the Department of Neurology, Medical University of Graz, Austria (2011-2017) were retrospectively analyzed. Details on the methods, including patient inclusion, clinical follow-up, and post-interventional MRI brain imaging, are given in the supplementary material and in the original paper^4^. Exclusion criteria were missing MRI, age <18 years, prior seizures or pre-existing brain lesions, and death within 24 hours. The ethics committees of Johannes Kepler University Linz and the Medical University of Graz approved the original study (Approval Nos. Linz: 1183/2020; Graz: 32-634 ex 19/20). This study was conducted in accordance with the ethical principles outlined in the Declaration of Helsinki.

### Quantification of damage in NT-informed structural networks

Binary lesion masks were embedded into previously published normative neurotransmitter (NT)-informed structural connectomes derived from PET receptor/transporter density maps and a population-averaged structural connectome^27,29,30^. In brief, NT-specific streamline weights, reflecting receptor and transporter densities at streamline endpoints, were normalized within each NT system to enable comparability across systems. Lesion-induced NT-network damage was then quantified as the normalized sum of weighted streamlines intersecting the lesion. In parallel, we quantified global structural disconnection as the total number of disconnected streamlines (SL). Additional technical details are provided in the Supplementary Methods and in recent methodological work ^29,31,32^. Please see Figure 1 for an illustration of the analysis pipeline.

**Figure 1.**
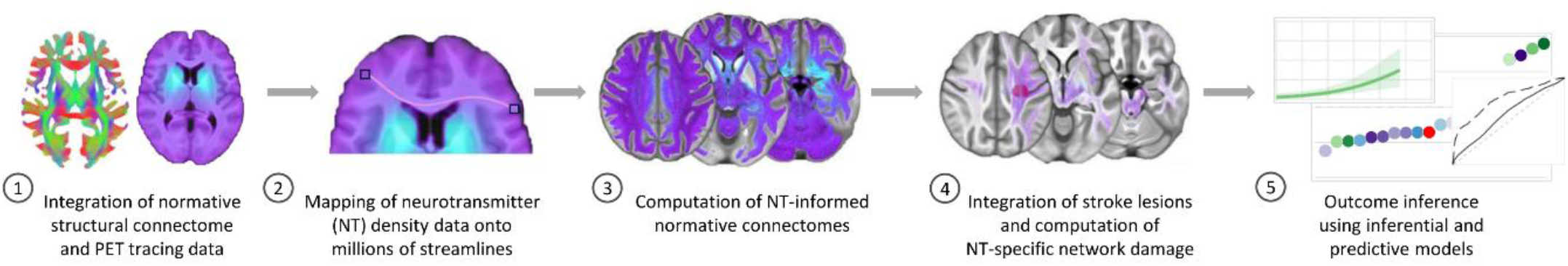
Analysis pipeline. **1**. Normative neurotransmitter (NT) receptor and transporter density maps derived from PET data were projected onto a population-averaged structural connectome. **2/3**. NT-specific normative connectomes were computed by weighting individual streamlines according to receptor/transporter densities at their endpoints. **4**. Lesion masks were then integrated to quantify NT-specific network damage by summing the weighted streamlines intersecting the lesion. This procedure yielded normalized NT-specific damage scores. (see https://github.com/phjkoch/NTDisconn) **5**. These were related to clinical outcomes using correlative modeling and inferential plus internally cross-validated resampling analyses.

### Statistical analysis

To assess the association between NT-informed network damage and PSE, we employed a multilevel analytical framework that explicitly separates feature extraction and inference from internal resampling-based model evaluation. This strategy was chosen to address the pronounced multicollinearity among NT-informed network damage measures and to distinguish explanatory associations from the performance of internally cross-validated models.

Partial least squares (PLS) regression was applied to identify informative predictors using variable importance in projection (VIP) scores^33-36^. Predictors exceeding the prespecified VIP threshold above 1 were subsequently entered into multivariable logistic regression models with PSE as the outcome, adjusting for age, sex, and the SeLECT score^37^. Model fit was evaluated relative to the covariate-only base model using ΔAIC and Nagelkerke’s pseudo-R^2^, with false-discovery rate correction applied across predictor-level tests.

To complement inferential analyses, we explored model performance under internal resampling using elastic-net–regularized logistic regression with repeated stratified 5-fold cross-validation.

Full models, including NT-informed damage measures and SL, were again compared with the covariate-only base model. Predictive performance was quantified on held-out folds using ROC-AUC and PR-AUC, and predictor contributions were summarized by selection frequency and standardized coefficient magnitude. Please see the Supplementary material for details on correlative modeling, inferential analyses, and internal resampling analyses.

## Results

### Cohort characteristics

The final cohort comprised 251 patients with acute large-vessel occlusion ischemic stroke treated with mechanical thrombectomy, of whom 26 patients (10.4%) developed post-stroke epilepsy (PSE) during follow-up. The overall mean age was 63.0 ± 14.6 years, with a comparable sex distribution across groups (48.6% female overall; 50.0% in the PSE group). Baseline stroke severity was moderate to severe, with a median admission NIHSS score of 14.0 (IQR 9.0-17.0), improving to a median follow-up NIHSS score of 4.0 (IQR 2.0-9.0). Patients who developed PSE tended to have higher NIHSS scores both at admission (median 14.5 [IQR 11.0-19.8]) and at follow-up (median 7.5 [IQR 4.2-13.8]) compared with patients without PSE (median 14.0 [IQR 9.0-17.0] and 4.0 [IQR 2.0-8.0], respectively). Mean lesion volume on postinterventional MRI was substantially larger in patients with PSE (204.6 ± 140.3 mL) than in those without PSE (80.1 ± 88.5 mL). Intravenous thrombolysis was administered in 68.5% of patients overall, with similar rates in patients with and without PSE. Successful reperfusion was frequent, with TICI 2b-3 achieved in 92.0% of cases and did not differ between groups. Stroke etiology was predominantly cardioembolic (45.0%), followed by large-artery atherosclerosis (21.1%) and other or unknown causes (33.9%), with a comparable distribution between PSE and non-PSE patients. Early seizures within the first week after stroke were uncommon overall (1.6%), but occurred more frequently among patients who later developed PSE (11.5%) than among those without PSE (0.4%). Consistent with this finding, the median SeLECT score was higher in patients with PSE (4.0 [IQR 4.0-5.0]) than in those without PSE (4.0 [IQR 3.0-5.0]). Regarding antidepressant medication, 37.1% of patients received selective serotonin reuptake inhibitors (SSRIs), 6.8% serotonin-norepinephrine reuptake inhibitors (SNRIs), and 40.2% other antidepressants. Antidepressant use tended to be more frequent among patients with PSE than among those without, particularly regarding SSRIs and other antidepressants (Table 1). Please see Figure 2 for the distribution of stroke lesions and structural disconnection patterns in the patient groups.

**Table 1.** Clinical and paraclinical data. Data are presented as mean ± SD for continuous variables, as median ± SD for ordinal variables (highlighted by asterisks), and n (%) for categorical variables. Abbreviations: FU = follow-up; IQR = interquartile range; NIHSS = National Institutes of Health Stroke Scale; PSE = post-stroke epilepsy; SD = standard deviation; SeLECT = Severity of stroke, Large-artery atherosclerotic etiology, Early seizure, Cortical involvement, Territory of middle cerebral artery; SNRI = serotonin-norepinephrine reuptake inhibitor; SSRI = selective serotonin reuptake inhibitor; TICI = Thrombolysis in Cerebral Infarction.

| Category | Variable | Unit/Level | ALL (N=251) | No PSE (N=225) | PSE (N=26) |
| --- | --- | --- | --- | --- | --- |
| Demographic variables | Age | Years | $63.0 \pm 14.6$ | $63.4 \pm 14.7$ | $59.4 \pm 14.1$ |
|  | Sex | Female | 122 (48.6%) | 109 (48.4%) | 13 (50%) |
|  |  | Male | 129 (51.4%) | 116 (51.6%) | 13 (50%) |
| Stroke characteristics | NIHSS* | - | 14.0 (9.0-17.0) | 14.0 (9.0-17.0) | 14.5 (11.0-19.8) |
|  | NIHSS<sub>FU</sub>* | - | 4.0 (IQR 2.0-9.0) | 4.0 (IQR 2.0-8.0) | 7.5 (IQR 4.2-13.8) |
| | Lesion volume<sub>FU</sub> | mL | $92.9 \pm 102.1$ | $80.1 \pm 88.5$ | $204.6 \pm 140.3$ |
|  | Etiology | Large artery atherosclerosis | 53 (21.1%) | 49 (21.8%) | 4 (15.4%) |
|  |  | Cardioembolic | 113 (45%) | 102 (45.3%) | 11 (42.3%) |
|  |  | Other | 85 (33.9%) | 74 (32.9%) | 11 (42.3%) |
| Treatment and Recanalization | Thrombolysis | no | 79 (31.5%) | 70 (31.1%) | 9 (34.6%) |
|  |  | yes | 172 (68.5%) | 155 (68.9%) | 17 (65.4%) |
|  | TICI-score | 0 | 15 (6%) | 13 (5.8%) | 2 (7.7%) |
|  |  | 1 | 1 (0.4%) | 1 (0.4%) | - |
|  |  | 2a | 4 (1.6%) | 4 (1.8%) | - |
|  |  | 2b | 51 (20.3%) | 45 (20%) | 6 (23.1%) |
|  |  | 2c | 34 (13.5%) | 31 (13.8%) | 3 (11.5%) |
|  |  | 3 | 146 (58.2%) | 131 (58.2%) | 15 (57.7%) |
| Prognostic factors | Early Seizures | no | 247 (98.4%) | 224 (99.6%) | 23 (88.5%) |
|  |  | yes | 4 (1.6%) | 1 (0.4%) | 3 (11.5%) |
|  | SeLECT* | - | 4.0 (3.0-5.0) | 4.0 (3.0-5.0) | 4.0 (4.0-5.0) |
| Antidepressants | SNRI | no | 234 (93.2%) | 211 (93.8%) | 23 (88.5%) |
|  |  | yes | 17 (6.8%) | 14 (6.2%) | 3 (11.5%) |
|  | SSRI | no | 158 (62.9%) | 144 (64%) | 14 (53.8%) |
|  |  | yes | 93 (37.1%) | 81 (36%) | 12 (46.2%) |
|  | Other | no | 150 (59.8%) | 136 (60.4%) | 14 (53.8%) |
|  |  | yes | 101 (40.2%) | 89 (39.6%) | 12 (46.2%) |

**Figure 2.**
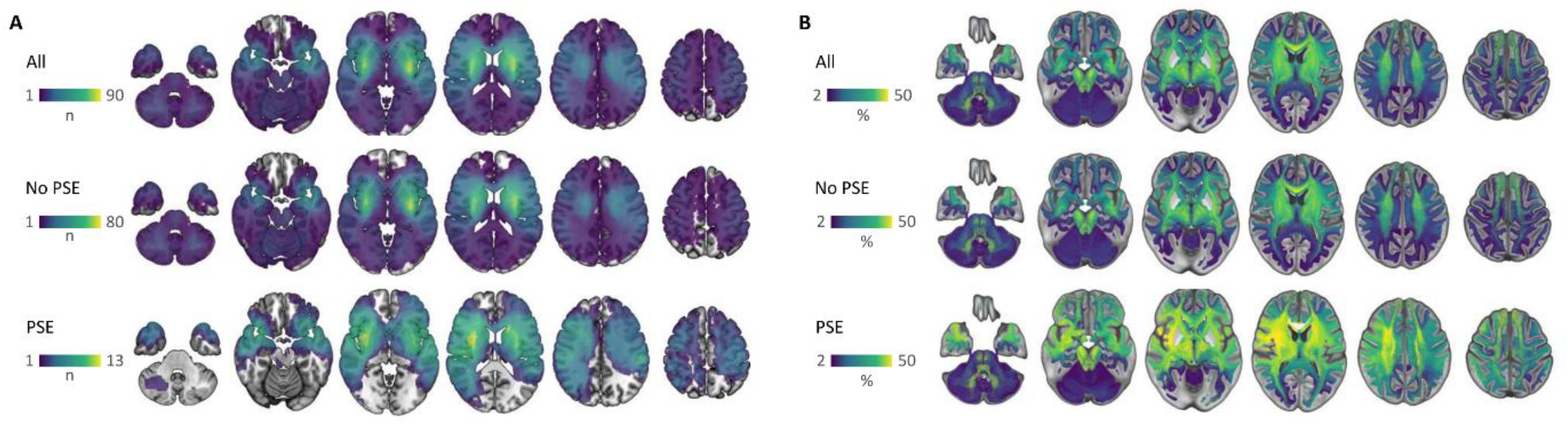
Lesion distribution and structural disconnection patterns in patients with and without PSE. **A** Group-level lesion overlay maps in MNI space shown for the full sample (All), patients without post-stroke epilepsy (No PSE), and patients with post-stroke epilepsy (PSE). Color intensity indicates the number of patients with lesion involvement in each voxel (n). B Group-level structural disconnection (SL) overlay maps derived from disconnectome analysis, displayed in MNI space for the same subgroups (All, No PSE, PSE). Color intensity represents the percentage of disconnected streamlines, averaged across individuals at the group level (%).

### Distribution of neurotransmitter-informed network disconnection

Neurotransmitter-informed network damage scores showed systematic differences across NT systems (Fig. 3A). A one-way analysis of variance revealed a significant main effect of NT system on network damage (F_18,4750_ = 7.99, *P*<2×10^−16^), indicating a non-uniform neurochemical vulnerability profile in LVO stroke. Post-hoc Tukey tests demonstrated that serotonergic networks, particularly those characterized by the serotonin transporter (5-HTT) and 5-HT_4_ receptors, exhibited significantly greater mean network damage than several other NT systems, including GABAergic, glutamatergic, and dopaminergic networks (all adjusted *P*<0.001).

**Figure 3.**
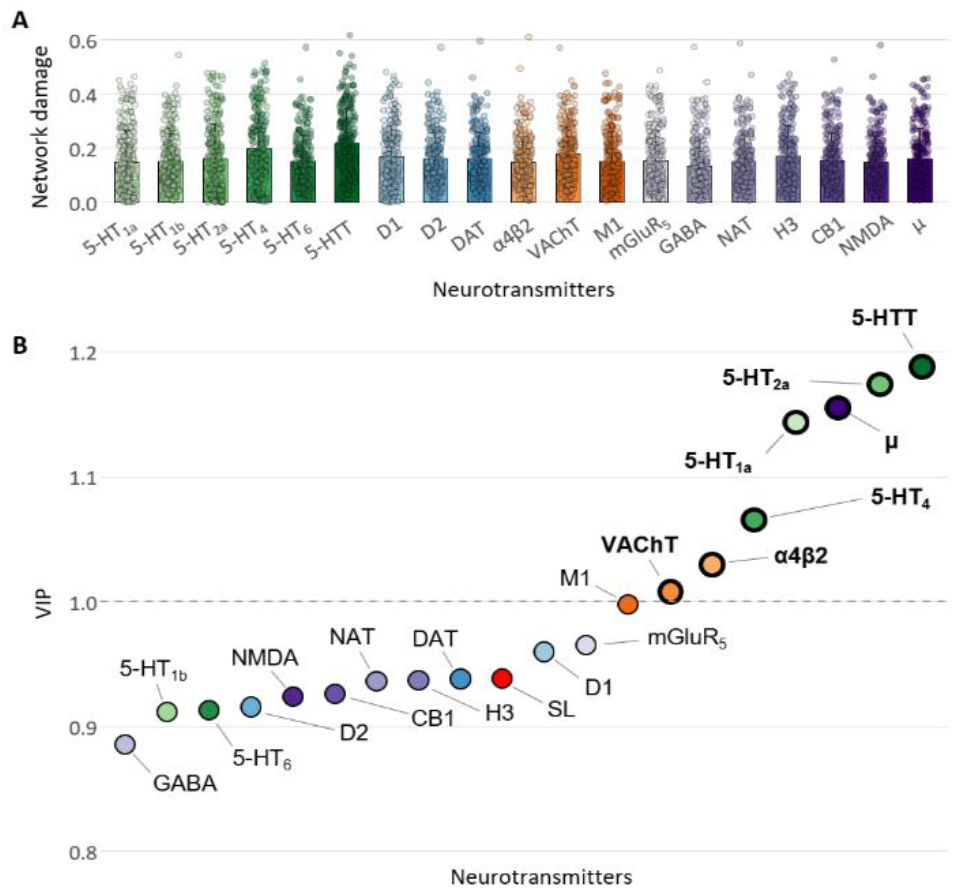
Neurotransmitter-informed network damage and PLS-based predictor selection. **A** Network damage scores (0-1) are plotted for the 19 neurotransmitter receptor and transporter networks (see Suppl. Table 1). Informative predictors for the inference of PSE (**B**) were selected using partial least squares (PLS) regression and cumulative variable importance in projection (VIP) scores across three latent components, aiming to stabilize variable weights and capture distributed covariance patterns in multivariate data. Informative predictors were defined as VIP>1 (points with bold circles). These underwent further GLM analysis to obtain adjusted interpretations. For visualization, data were grouped according to the NT system, with green representing serotonin, blue representing dopamine, orange representing acetylcholine, and purple representing other

In contrast, GABA-informed networks showed the lowest average damage levels. Ranking NT systems by mean damage revealed the highest values for 5-HTT (mean 0.22 ± 0.13) and 5-HT_4_ (0.20 ± 0.13), followed by VAChT, H3, and D1. These findings indicate a preferential involvement of serotonergic and selected cholinergic, histaminergic, and dopaminergic networks in LVO stroke (Fig. 3A). To assess group differences between patients without and with PSE, neurotransmitter-specific network damage scores were compared using NT-wise Welch t-tests with false discovery rate (FDR) correction. Across all neurotransmitter systems, patients with PSE exhibited significantly higher network damage than patients without PSE (all FDR-corrected *P*<0.001). The largest absolute differences were observed for serotonergic networks, including 5-HT_1a_, 5-HT_2a_, 5-HT_4_, 5-HT_6_, and the serotonin transporter (5-HTT), where mean network damage in the PSE group exceeded that of patients without PSE by approximately 0.10-0.14 units (Supplementary Table 1). Collectively, these findings indicate a global increase in the burden of neurotransmitter-specific network disconnection in patients with PSE, superimposed on a baseline pattern of preferential serotonergic vulnerability observed across the entire cohort.

### PLS regression for variable selection

To identify the most informative predictors of PSE, NT-specific damage scores and the global structural disconnection measure (SL) were entered into a PLS regression model. Variable importance was assessed using cumulative VIP scores (Fig. 3B). Seven NT systems were identified as informative: serotonergic 5-HTT, 5-HT_1a_, 5-HT_2a_, 5-HT_4_, μ-opioid, and cholinergic VAChT, and α4β2 nicotinic receptors. The global structural disconnection measure (SL) did not reach the VIP threshold. Notably, the majority of selected predictors belonged to the serotonergic system, highlighting its dominant contribution to the multivariate association with PSE. In contrast, GABAergic and glutamatergic markers did not reach the VIP threshold, indicating a comparatively lower contribution after accounting for shared variance across NT systems.

### Adjusted modeling for validation and interpretation

To validate and interpret the associations identified by PLS, each selected predictor was entered into generalized linear models with PSE as the binary outcome, adjusting for the base variables age, sex, and the SeLECT score (Table 2, Figure 4). All serotonergic markers identified by PLS showed significant independent associations with PSE. The strongest effects were observed for 5-HT_1a_ (OR 2.09, 95% CI 1.39-3.26, *P*_FDR_ = 0.002) and 5-HT_2a_ (OR 1.96, 95% CI 1.35-2.94, *P*_FDR_ = 0.002). Here, a 10% increase in NT-network damage translated to a two-fold increase in PSE risk. Similarly, 5-HT_4_ (OR 1.92, 95% CI 1.29-2.95) and 5-HTT (OR 1.61, 95% CI 1.08-2.44) were significantly associated with PSE. Beyond the serotonergic system, µ-opioid receptor-informed network damage was associated with increased PSE risk (OR 2.22, 95% CI 1.42-3.56, *P*_FDR_ = 0.002), as were cholinergic markers VAChT (OR 1.93, 95% CI 1.20-3.20) and α4β2 (OR 1.86, 95% CI 1.17-3.04). The global structural disconnection measure (SL) was also significantly associated with PSE (OR 1.93, 95% CI 1.21-3.14, *P*_FDR_ = 0.010). All predictors significantly increased pseudo-R^2^ by up to 10% in 5-HT_1a_, 5-HT_2a_, and µ, compared to the base model that explained 11% of variance in PSE with statistical significance for the factor SeLECT (OR 1.76, 1.28-2.47, P<0.001), but not age or sex. Notably, the NT-informed models for 5-HT_1a_, 5-HT_2a_, 5-HT_4_, and µ-opioid receptor demonstrated significantly greater improvements in model fit than SL (ΔAIC difference > 2), indicating that NT-specific network damage provided explanatory value beyond unspecific structural disconnection.

**Table 2.** Association between neurotransmitter-informed network damage and PSE. Generalized linear models examine the association between raw NT/SL damage scores and PSE. Models are adjusted for age, sex, and SeLECT score. Statistical significance is reported for each predictor within each model; a false discovery rate (FDR) correction was applied to account for 8 tests (*P*_FDR_). Odds ratios (OR) are given with 95% confidence intervals for a 10% increase in NT/SL damage. *mark statistical significance. The changes in AIC and R^2^ are computed relative to the base model, which includes only age, sex, and the SeLECT score. A change of >2 in AIC is considered a significant improvement in the model.

| Neurotransmitter / SL | OR (95% CI) | $P_{FDR}$ | $\Delta AIC$ | $\Delta R^2$ |
| --- | --- | --- | --- | --- |
| 5-HT<sub>1a</sub> | 2.09 (1.39-3.26) | 0.002** | 10.9 | 0.097 |
| 5-HT<sub>2a</sub> | 1.96 (1.35-2.94) | 0.002** | 10.7 | 0.096 |
| $\mu$ | 2.22 (1.42-3.56) | 0.002** | 10.7 | 0.096 |
| 5-HT<sub>4</sub> | 1.92 (1.29-2.95) | 0.004** | 8.9 | 0.082 |
| VACHT | 1.93 (1.2-3.2) | 0.010* | 5.4 | 0.057 |
| $\alpha 4\beta 2$ | 1.86 (1.17-3.04) | 0.011* | 4.8 | 0.052 |
| 5-HTT | 1.61 (1.08-2.44) | 0.021* | 3.6 | 0.043 |
| SL | 1.93 (1.21-3.14) | 0.010* | 5.65 | 0.058 |

**Figure 4.**
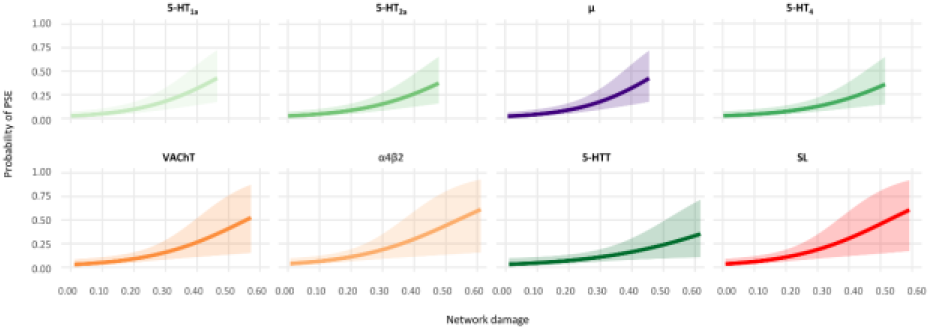
Adjusted effects of neurotransmitter-informed network damage on PSE. Adjusted marginal effects from generalized linear models illustrating the association between raw NT/SL damage scores and PSE for winning models from Tab. 2. Solid lines represent model-based predicted probabilities across the observed NT/SL damage range, while shaded ribbons indicate 95% confidence intervals. The NT-informed models for 5-HT_1a_, 5-HT_2a_, 5-HT_4_, and µ-opioid receptor (upper row) demonstrated significantly greater improvements in model fit than the base model (see text).

### Sensitivity analyses

To assess the robustness of the identified associations, we performed a series of stepwise sensitivity analyses in which each winning predictor with a VIP score > 1 was re-evaluated after the individual inclusion of additional clinical covariates beyond the base model (including age, sex, and SeLECT score). Specifically, models were successively adjusted for post-stroke neurological deficit (NIHSS_FU_), antidepressant treatment (SSRI, SNRI, or other antidepressants), and lesion volume after the acute treatment. To avoid collinearity between lesion size and network damage measures, lesion volume was included after a cube-root transformation to improve its distribution and as the residualized component after regression on the respective predictor, in line with previous studies ^38,39^. Across all sensitivity analyses, all winning predictors remained statistically significant (*P* < 0.05) after the addition of each individual covariate. This applied consistently to all serotonergic markers (5-HT_1a_, 5-HT_2a_, 5-HT_4_, 5-HTT), the µ-informed network, cholinergic markers (VAChT and α4β2), and SL. Notably, adjustment for antidepressant medication did not attenuate the observed associations, including those involving serotonergic network damage. Likewise, controlling for post-stroke neurological deficit and lesion volume did not materially alter the significance of any predictor. These findings indicate that the associations between NT-informed network damage and PSE are robust to potential confounding by clinical severity, treatment effects, and lesion size.

### Internal cross-validated model performance

While the PLS-GLM framework identified robust and biologically interpretable associations between NT-informed network damage and PSE, it was not designed to assess model performance under regularization and internal resampling. We therefore examined whether NT-informed network damage retained incremental signal beyond the SeLECT score, age, and sex using elastic-net–regularized logistic regression with repeated stratified cross-validation. Across 50× repeated stratified 5-fold cross-validation, the full model, including all NT-informed network damage measures and global structural disconnection (SL), showed higher discrimination than the clinical base model (Fig. 5A; AUC mean ± SD: base 0.64±0.11, full 0.74±0.11, P<0.001). Precision-recall analysis provided complementary information under class imbalance and likewise favored the full model (Fig. 5B). Although absolute precision values remained modest, the full model consistently achieved higher precision across a broad range of recall values than the base model. This indicates that NT-informed network damage contributes clinically relevant information for identifying individuals at increased PSE risk.

**Figure 5.**
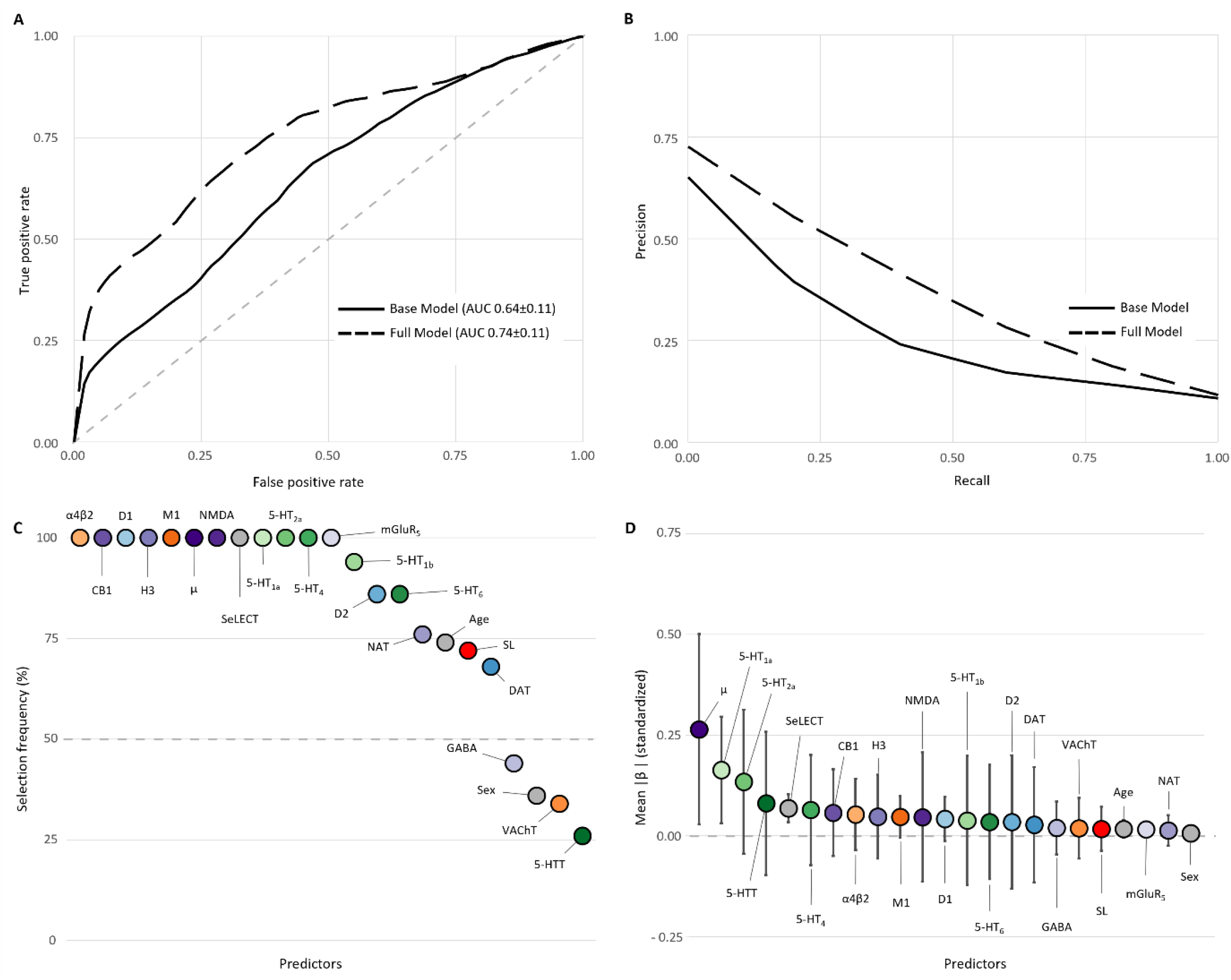
Internal cross-validated model performance. **A** Mean receiver operating characteristic (ROC) curve comparing a base elastic-net model (solid) with a full elastic-net model (dashed), including NT-informed network damage measures and structural lesion damage, evaluated by repeated stratified cross-validation. ROC-AUC is reported as mean ± SD across held-out test folds. **B** Corresponding mean precision-recall curves, highlighting improved performance of the full model under class imbalance (summarized across held-out test folds). **C** Selection frequency in % of individual predictors across repeats for the full model, indicating stability (dashed line denotes 50% selection probability). **D** Mean absolute standardized regression coefficients (|β| ± SD) for predictors in the full model at the selected penalty, summarizing relative effect magnitude across resampling iterations.

To characterize how individual predictors contributed within this regularized resampling framework, standardized elastic-net coefficients were summarized across repeats. NT-informed predictors differed markedly in both selection stability and effect magnitude (Fig. 5C, D). Serotonergic network damage measures, particularly 5-HT_1a_ and 5-HT_2a_, showed high selection frequencies and consistent coefficient directions across resampling iterations. The µ-opioid– informed network emerged as the strongest single predictor, ranking highest in mean absolute standardized coefficient magnitude and showing maximal selection stability. In contrast, SL was selected less consistently and contributed smaller effect sizes, despite its significant association with PSE in inferential models. Age and sex contributed minimally and were rarely retained by the regularized models. The SeLECT score showed modest and unstable selection across resampling iterations, suggesting that NT-informed network damage retained information not consistently represented by the clinical covariates alone. In a sensitivity analysis, lesion volume was added to the model; although it was selected in all iterations, its standardized effect size was negligible, indicating limited additional signal beyond NT-informed network damage (not shown).

### Convergence between inference and internal resampling analyses

Taken together, inferential and internal resampling analyses converged on a consistent core signature involving serotonergic and µ-opioid networks. NT systems identified as informative by PLS and validated in adjusted GLMs also emerged as the most stable contributors across cross-validation iterations.

In contrast, global structural disconnection and cholinergic markers contributed less consistently, underscoring the neurochemical specificity of the observed signal. Collectively, these findings indicate that NT-informed network damage not only explains inter-individual variability in PSE risk but also retains a stable signal under regularized internal resampling.

## Discussion

In this multicenter cohort of thrombectomy-treated large-vessel occlusion stroke patients, lesion-induced disruption of neurotransmitter-informed structural networks was associated with post-stroke epilepsy. Across complementary analytical levels, the results converged on a robust core signature involving damage to networks related to serotonergic 5-HT_1a_ and 5-HT_2a_ and µ-opioid systems.

These predictors showed the strongest effect estimates in adjusted models, provided the largest improvements in model fit, explained variance beyond the clinical baseline, remained significant in sensitivity analyses adjusting for post-stroke deficit, antidepressant exposure, and lesion volume, and emerged as the most stable contributors across internal cross-validation analyses. Other PLS-selected predictors, including cholinergic VAChT and α4β2, appeared less central. Together, these findings suggest that the neurochemical fingerprint of disconnected networks contributes to epileptogenesis after severe stroke.

### Serotonergic network disruption and epilepsy

Serotonin is a state-dependent neuromodulator that shapes cortical and limbic excitability, interneuron recruitment, and network synchronization. Across preclinical epilepsy models, reduced serotonergic tone generally increases seizure susceptibility, whereas serotonergic enhancement can raise seizure thresholds, albeit in a receptor-, region-, and dose-dependent manner^18^. In line with this, serotonin depletion increased seizure susceptibility and worsened neuropathological outcomes in a kainate model of epilepsy, and stimulation of 5-HT_1a_ receptors increased seizure thresholds in a picrotoxin convulsion model^20,40^. These experimental data provide a plausible framework for our observation that greater disruption of serotonergic structural networks, as captured by lesions that disconnect regions with high serotonergic receptor/transporter densities, is associated with higher PSE risk.

A potential concern is that some clinical literature links selective serotonin reuptake inhibitor (SSRI) treatment to a higher risk of seizures after stroke, including meta-analytic evidence of increased seizure risk with SSRIs versus placebo and large randomized fluoxetine trials designed for post-stroke recovery^41-43^. These observations do not contradict the present results for several reasons. First, SSRI prescription after stroke is prone to confounding by indication (depression and worse clinical status) and to time-varying confounding, both of which may inflate apparent seizure risk in nonrandomized settings. Second, trial data indicate that fluoxetine can modestly increase seizures at the population level, yet this pharmacological effect is not equivalent to that of structural serotonergic network injury. Importantly, in our data, serotonergic network damage was linked to PSE independently of documented antidepressant exposure. Third, preclinical work suggests that SSRIs can have bidirectional effects on epileptogenesis depending on brain state and model: fluoxetine accelerated epileptogenesis and increased seizure burden in a rat model of acquired epilepsy, consistent with the possibility that enhancing serotonin signaling in an injured, plastic brain can sometimes promote maladaptive network reorganization^44^. Conversely, reviews of antidepressants in epilepsy emphasize that modern antidepressants are often not strongly proconvulsant at therapeutic doses and that seizure risk may relate to underlying vulnerability rather than the drug per se^45^.

The strongest associations involved networks characterized by distributions of 5-HT_1a_, 5-HT_2a_, 5-HT_4_, and the serotonin transporter (5-HTT). 5-HT_1a_ signaling can exert anticonvulsant effects via hyperpolarization and modulation of inhibitory interneurons; accordingly, stimulation of 5-HT_1a_ receptors increases seizure thresholds in rodent models^40^. 5-HT_2_-family signaling, particularly involving 5-HT_2c_ receptors, has been implicated in the regulation of cortical excitation and seizure propagation in a context-dependent manner ^18,46^. Recent preclinical work further supports a critical role of 5-HT_4_ receptors in serotonergic seizure protection. In a DBA/1 mouse model of SUDEP, selective antagonism of 5-HT_4_ receptors was the only receptor-level manipulation that reversed fenfluramine’s ability to block seizures and seizure-induced respiratory arrest, whereas pharmacological activation of 5-HT_4_ receptors reduced seizure-induced respiratory arrest and seizures ^47^. More broadly, a recent critical review emphasizes that modulation of distinct 5-HT receptor subtypes (including 5-HT_1_, 5-HT_2_, and 5-HT_4_) can yield divergent, model-dependent effects on epileptic activity and highlights serotonergic modulation, especially receptor-selective approaches, as a promising treatment avenue in selected epilepsy syndromes ^46^. Taken together, the clustering of 5-HT_1a_, 5-HT_2a_, 5-HT_4_, and 5-HTT among the most informative predictors suggests that lesions that disconnect serotonergic projection architecture and serotonergically enriched target regions may compromise stabilizing neuromodulatory control and thereby lower the seizure threshold after stroke ^18^.

### Beyond serotonin: opioid and cholinergic network damage

µ-opioid receptor-informed network damage was the most robust non-serotonergic marker of PSE across inference and prediction. In animal models, opioid signaling has been linked to the pathophysiology of hyperexcitability and early post-stroke seizures^26^. Following cerebral ischemia, µ-opioid receptor expression is upregulated, particularly in hippocampal granule cells, a response thought to reflect increased neuronal excitability. This induction may serve a compensatory role by modulating excessive glutamatergic excitation through inhibitory opioid mechanisms^48^. Further, animal models and human studies have demonstrated increased µ-receptor density in epileptogenic regions, including the hippocampus, thalamus, and cortex, after seizures or ischemic injury. µ-receptor activation can exert proconvulsant effects by lowering seizure thresholds and facilitating epileptic activity^49,50^. In parallel, evidence suggests that pharmacological µ-receptor blockade with antagonists such as naltrexone may attenuate epileptogenic processes following hypoxic-ischemic injury^26^. Thus, these findings indicate a bidirectional role of µ-opioid receptors in post-stroke epileptogenesis. µ induction and activation after stroke may exert either proconvulsive or compensatory inhibitory effects on neuronal excitability, depending on regional distribution, timing, and interactions with other neurochemical systems.

Cholinergic network damage (VAChT and α4β2) was also associated with an increased risk of PSE. Acetylcholine is a powerful regulator of cortical state and synchronization; experimental studies show that cholinergic activation can elicit seizure-like activity and that cholinergic signaling is altered in chronic epilepsy models and human epileptogenic tissue^21-23^. However, their smaller AIC/ΔR^2^ gains and weaker stability in elastic-net selection suggest that cholinergic effects may be context-dependent or reflect shared variance with more dominant NT patterns rather than an independent signal of equal strength. This does not rule out a mechanistic role of cholinergic pathways, but it indicates that, in the present dataset, cholinergic predictors contribute less robustly to generalizable risk stratification than the core serotonergic and opioid markers.

### Neurotransmitter-informed vulnerability beyond global disconnection

Global disconnection (SL) was significantly associated with PSE in inferential models, supporting network-level conceptualizations of lesion-related epilepsy^10^. However, the fact that the core NT-informed predictors provided larger improvements in model fit than SL and were preferentially retained across internal cross-validation analyses indicates that epileptogenesis after severe stroke relates not only to the extent of disconnection but also to its neurochemical specificity. This extends prior connectome-based work in PSE by adding a mechanistically interpretable neurochemical layer that may help explain why lesions of similar size or location differ in epileptogenic potential.

### Clinical implications and limitations

Neurotransmitter-informed network damage may complement established clinical risk prediction models, such as SeLECT, for stratifying post-stroke epilepsy after mechanical thrombectomy. Because the observed associations remained significant after individual adjustment for antidepressant exposure, post-stroke neurological deficit, and lesion volume, they are unlikely to merely reflect treatment effects or global stroke severity. Several limitations should be considered. First, NT-informed network damage was derived from normative PET receptor maps and a population-averaged structural connectome, rather than from individual receptor densities or patient-specific connectivity, which may attenuate subject-level specificity. Second, outcome ascertainment was retrospective, and the number of PSE events was moderate, reflecting the relatively low incidence of epilepsy in this clinical population. To address this, model performance was examined using repeated stratified internal cross-validation with regularization, which stabilizes performance estimates in the presence of class imbalance but does not replace external validation. Consequently, these findings should be interpreted as internal resampling results rather than definitive clinical predictive performance. Third, although elastic-net modeling identified a subset of NT systems as stable contributors across internal resampling analyses, most consistently serotonergic (5-HT_1a_, 5-HT_2a_) and µ-opioid-informed networks, these results do not imply causal mechanisms and may still be influenced by residual collinearity across neurochemical systems. Notably, cholinergic markers that showed weaker inferential support in PLS-GLM analyses also exhibited lower stability of selection in the resampling analyses, underscoring the importance of converging evidence across analytical frameworks. Finally, the cohort was restricted to patients with MRI availability after mechanical thrombectomy, which may limit generalizability to the broader stroke population. Future prospective studies with larger event numbers and independent validation cohorts should evaluate combined clinical-neurochemical models and test whether NT-informed network vulnerability can inform seizure surveillance after stroke.

## Data Availability

Data available upon reasonable request

## Acknowledgments

The authors acknowledge the use of artificial intelligence–based tools to support this work (ChatGPT version 5.2, OpenAI, San Francisco, CA, USA; and OpenEvidence, OpenEvidence Inc.). AI assistance was employed for (i) code troubleshooting for statistical analyses and data visualization, (ii) targeted literature search and synthesis, and (iii) language editing and structural refinement of the manuscript draft. All analyses, interpretations, and final manuscript content were critically reviewed, verified, and approved by the authors, who take full responsibility for the accuracy and integrity of the work. This manuscript has been posted as a preprint on www.medrxiv.org.

