## Supplementary Material for "Damage to serotonergic and opioid networks relates to post-stroke epilepsy after thrombectomy"

### Supplementary Methods

*Participants and lesion delineation*

Our final study cohort comprised 251 thrombectomy patients with a median long-term follow-up of
90 months (range 0-142). The study size was determined by all consecutive eligible patients meeting inclusion criteria during the study period; no formal sample size calculation was performed. Clinical follow-up in the Linz cohort was performed via a structured, validated telephone interview to detect seizures; if needed, close relatives, nursing staff, or general practitioners were contacted. An in-person neurological evaluation and an EEG followed any positive screening results. For the Graz cohort, follow-up was obtained from KAGes, the electronic documentation system covering all public hospitals in Styria, which enabled identification of any seizure-related visit or hospitalization. Post-interventional MRI (typically obtained on day 1) was assessed by three trained raters blinded to seizure outcomes. Stroke lesions were manually delineated on MRI using fluid-attenuated inversion recovery (FLAIR) scans acquired on 1.5 T or 3 T scanners. Lesion delineation followed a semi-automated outlining procedure with subsequent generation of binarized lesion masks[1].

*Quantification of damage in NT-informed structural networks*

To quantify lesion-induced damage in NT-informed structural networks, the binary lesion masks were integrated into normative NT-informed structural connectome data, as previously reported[2]. Figure 1 summarizes the analysis pipeline. In brief, 19 different normative NT receptor and transporter density maps, i.e., positron-emission tomography (PET) tracer maps for serotonergic, dopaminergic, cholinergic, glutamatergic, GABAergic, and noradrenergic, histaminergic, cannabinoid and opioid NT systems, were obtained from Hansen *et al.*[3]. Each PET tracer map was co-registered to an averaged Human Connectome Project-derived normative connectome with 2 million streamlines (SL)[4]. Each SL was assigned the product of specific NT densities in its endpoints, defined as the NT-specific streamline weight ($SL_{NTW}$). To allow for comparability between different NT systems, $SL_{NTW}$ data were normalized to the sum of weights across all SL within each NT map. Thus, 19 new normative NT-informed connectivity maps in MNI space are generated and are publicly available on GitHub (phjkoch/NTDisconn). Higher $SL_{NTW}$ values within these connectomes indicate an SL that connects areas with higher receptor and transporter NT densities, respectively. By integrating binary lesion masks into these connectivity maps, we collected all SL that passed through the lesion and summed their respective NT weights. Given the normalization step, this estimate of NT network damage (the sum of SL weights) ranges from 0 (0% damage) to 1 (100% damage). In addition to NT damage, we computed the sum of disconnected SL as a global measure of network damage (abbreviated as SL in the figures for simplicity).

*Correlative modeling and inferential analyses*

First, all NT-informed network damage scores and the unbiased measure of structural disconnection (SL), which are multicollinear by construction, were subjected to partial least squares (PLS) regression using the plsr function in R (version 4.4.1). Variable importance was quantified using Variable Importance in Projection (VIP) scores, reflecting each predictor's cumulative contribution across latent components. Predictors with VIP > 1 were considered informative. Although 10-fold cross-validation indicated that a single latent component minimized prediction error (root mean squared error of prediction, RMSEP), three latent components were retained for VIP computation. This decision followed established recommendations that slightly exceeding the predictive optimum improves the stability of variable weights and better captures distributed covariance structures in multivariate neuroimaging data[5-8]. Predictors exceeding the VIP threshold were subsequently entered into logistic regression models (GLMs) for inferential interpretation. In contrast to PLS, these models estimate the association between individual NT-informed network damage measures and PSE while adjusting for clinically relevant covariates, including age, sex, and the SeLECT score[9], a validated clinical risk score for post-stroke epilepsy. The SeLECT score is based on five routinely available clinical parameters: stroke severity (NIHSS at admission), large-artery atherosclerotic etiology, early seizures (within 7 days after stroke), cortical involvement, and middle cerebral artery (MCA) territory involvement. Predictor-level significance was assessed using z-tests, and model-level evidence was evaluated by comparing full models against covariate-only base models. To determine whether including a given NT-informed predictor or SL improved model fit, Akaike Information Criterion (AIC) values were compared, with $\Delta$AIC $\geq 2$ indicating a meaningful improvement. Changes in explained variance were quantified using Nagelkerke's pseudo-$R^2$, derived from differences in log-likelihood between full and intercept-only models. P-values were corrected for multiple comparisons using the false discovery rate (FDR). Odds ratios (ORs) were reported for a 10% increase in NT-informed or SL-related network damage. For comparability, SL-related models were additionally explored independently of their VIP results.

*Internal cross-validated model performance*

While the PLS-GLM framework provides stable feature selection and interpretable inference, association-based results do not directly assess model performance under regularization and internal resampling. We therefore complemented the inferential analyses with a regularized internal cross-validation framework using elastic-net logistic regression (R package glmnet). Elastic net regression is well-suited for correlated predictors because it can retain groups of related variables while controlling overfitting via regularization. Predictive performance was estimated using repeated stratified 5-fold cross-validation (50 repeats). For each repeat, folds were constructed to preserve the outcome prevalence within each fold. Models were trained on 4/5 of the data and evaluated on the held-out 1/5 test fold. To account for class imbalance, class-balanced observation weights were applied within each training fold (weights proportional to the inverse class frequency; controls weight = 1, cases up-weighted by n0/n1). Within each outer training set, the regularization parameter λ was optimized by internal cross-validation (5 folds, deviance loss), and predictions were generated at λ_min (minimum cross-validated deviance), with glmnet's internal predictor standardization enabled. Two predictive models were evaluated: (i) a base model including age, sex, and SeLECT; and (ii) a full model including the base variables plus all NT-informed network damage measures and SL. Predictive performance on held-out test folds was quantified using ROC-AUC and precision-recall AUC (PR-AUC). Mean ROC and PR curves were obtained by interpolating fold-wise curves onto fixed grids and averaging across all folds and repeats. To characterize the contributions of predictors within the predictive framework, regression coefficients from the full model were extracted at λ_min. In addition to glmnet-scale coefficients, we computed standardized coefficients per fold (β_std = β*SD_train with SD computed from the original-scale predictors in the corresponding training set), yielding effect sizes for a +1 SD increase in the training data. For each predictor, we summarized the selection frequency (the proportion of repeats in which the predictor had a nonzero coefficient in at least one fold) and the mean absolute standardized coefficient magnitude, along with its variability across repeats, providing complementary measures of effect size and stability across resampling iterations.

**Supplementary Tables**

| Neurotransmitter | ALL | No PSE | PSE | Mean difference | $P_{FDR}$ |
|---|---|---|---|---|---|
| $5\text{-HT}_{1a}$ | 0.15 ± 0.11 | 0.14 ± 0.10 | 0.26 ± 0.14 | 0.13 ± 0.11 | 0.000*** |
| $5\text{-HT}_{1b}$ | 0.15 ± 0.10 | 0.14 ± 0.09 | 0.24 ± 0.11 | 0.10 ± 0.09 | 0.000*** |
| $5\text{-HT}_{2a}$ | 0.16 ± 0.13 | 0.15 ± 0.12 | 0.29 ± 0.15 | 0.14 ± 0.12 | 0.000*** |
| $5\text{-HT}_{4}$ | 0.20 ± 0.13 | 0.18 ± 0.12 | 0.32 ± 0.14 | 0.13 ± 0.12 | 0.000*** |
| $5\text{-HT}_{6}$ | 0.15 ± 0.10 | 0.14 ± 0.09 | 0.24 ± 0.11 | 0.10 ± 0.09 | 0.000*** |
| 5-HTT | 0.22 ± 0.13 | 0.21 ± 0.12 | 0.32 ± 0.14 | 0.11 ± 0.12 | 0.001*** |
| D1 | 0.17 ± 0.11 | 0.16 ± 0.11 | 0.27 ± 0.13 | 0.12 ± 0.11 | 0.000*** |
| D2 | 0.16 ± 0.10 | 0.15 ± 0.09 | 0.25 ± 0.11 | 0.10 ± 0.09 | 0.000*** |
| DAT | 0.16 ± 0.10 | 0.15 ± 0.09 | 0.25 ± 0.10 | 0.10 ± 0.09 | 0.000*** |
| α4β2 | 0.15 ± 0.09 | 0.14 ± 0.09 | 0.23 ± 0.10 | 0.09 ± 0.09 | 0.000*** |
| VAChT | 0.18 ± 0.10 | 0.17 ± 0.10 | 0.26 ± 0.11 | 0.10 ± 0.10 | 0.000*** |
| $\text{mGluR}_5$ | 0.16 ± 0.11 | 0.14 ± 0.10 | 0.26 ± 0.13 | 0.12 ± 0.10 | 0.000*** |
| GABA | 0.13 ± 0.10 | 0.12 ± 0.09 | 0.22 ± 0.11 | 0.10 ± 0.09 | 0.000*** |
| NAT | 0.15 ± 0.10 | 0.14 ± 0.09 | 0.24 ± 0.10 | 0.09 ± 0.09 | 0.000*** |
| H3 | 0.17 ± 0.11 | 0.16 ± 0.10 | 0.28 ± 0.12 | 0.12 ± 0.10 | 0.000*** |
| M1 | 0.15 ± 0.11 | 0.14 ± 0.10 | 0.26 ± 0.13 | 0.12 ± 0.10 | 0.000*** |
| CB1 | 0.15 ± 0.10 | 0.14 ± 0.09 | 0.25 ± 0.11 | 0.11 ± 0.10 | 0.000*** |
| NMDA | 0.15 ± 0.10 | 0.14 ± 0.09 | 0.23 ± 0.11 | 0.10 ± 0.09 | 0.000*** |
| μ | 0.16 ± 0.11 | 0.15 ± 0.10 | 0.27 ± 0.12 | 0.12 ± 0.10 | 0.000*** |

**SOM Tab. 1 | NT damage scores.** Neurotransmitter (NT)-informed network damage scores (range 0–1) are reported as mean ± standard deviation for the entire cohort (ALL), patients without PSE (No PSE), and patients with PSE (PSE). The mean difference represents the absolute difference in network damage between the PSE and No PSE groups (PSE - No PSE), expressed as mean difference ± pooled SD. Group comparisons were performed using NT-wise Welch two-sample t-tests, with false discovery rate (FDR) correction applied across all NT systems. $P_{FDR}$ denotes FDR-adjusted P values; *** indicates $P_{FDR}$ < 0.001. Neurotransmitters are ordered according to their appearance in Fig. 1A.
